# COVID-19 advocacy bias in the *BMJ*: meta-research evaluation

**DOI:** 10.1101/2024.06.12.24308823

**Authors:** Kasper P. Kepp, Ioana Alina Cristea, Taulant Muka, John P.A. Ioannidis

## Abstract

**Objectives:** During the COVID-19 pandemic, *BMJ*, the premier journal on evidence-based medicine worldwide, published many views by advocates of specific COVID-19 policies. We aimed to evaluate the presence and potential bias of this advocacy.

**Design and Methods:** Scopus was searched for items published until April 13, 2024 on “COVID-19 OR SARS-CoV-2”. *BMJ* publication numbers and types before (2016−2019) and during (2020−2023) the pandemic were compared for a group of advocates favoring aggressive measures (leaders of both the Independent Scientific Advisory Group for Emergencies (indieSAGE) and the Vaccines-Plus initiative) and four control groups: leading members of the governmental Scientific Advisory Group for Emergencies (SAGE), UK-based key signatories of the Great Barrington Declaration (GBD) (favoring more restricted measures), highly-cited UK scientists, and UK scientists who published the highest number of COVID-19-related papers in the entire scientific literature (n=16 in each group).

**Results:** 122 authors published more than 5 COVID-19-related items each in *BMJ*. Of those, 18 were leading members/signatories of aggressive measures advocacy groups publishing 231 COVID-19 related BMJ documents, 53 were editors/journalists, and 51 scientists were not identified as associated with any advocacy. Of 41 authors with >10 publications in *BMJ*, 8 were scientists advocating for aggressive measures, 7 were editors, 23 were journalists, and only 3 were non-advocate scientists. Some aggressive measures advocates already had strong *BMJ* presence pre-pandemic. During pandemic years, the studied indieSAGE/Vaccines-Plus advocates outperformed in *BMJ* presence leading SAGE members by 16.0-fold, UK-based GBD advocates by 64.2-fold, the most-cited scientists by 16.0-fold, and the authors who published most COVID-19 papers overall by 10.7-fold. The difference was driven mainly by short opinion pieces and analyses.

**Conclusions:** *BMJ* appears to have favored and massively promoted specific COVID-19 advocacy views during the pandemic, thereby strongly biasing the scientific picture on COVID-19.

**Summary box:** *Section 1: What is already known on this topic:*

- Advocacy is intensely debated for its merits to science and policy.
- Many journals increasingly publish pieces by advocates and it is thus important to understand the nature, scale and impact of this phenomenon.

*Section 2: What this study adds:*

- This study provides a detailed quantitative assessment of journal-promoted advocacy, focusing on the world’s premier evidence-based medical journal, the *BMJ*.
- We show that *BMJ* had massive bias towards specific COVID-19-related advocacy favoring aggressive measures.
- Our study reveals a need for editorial guidelines on journal-promoted advocacy.

## INTRODUCTION

Summary box

Science ideally develops conclusions from systematic evidence and balanced analysis of risks, intervention benefits and harms, and uncertainties.^1,2^ In contrast, advocacy groups lobby for specific policies, often in unilateral fashion not reflecting the full complexity of the issues involved. Advocacy has an important mission in raising awareness of critical needs. However, it may also be biased towards special ideological or financial interests that could sometimes harm society by unbalanced resource allocation.^3,4^

Leading medical and scientific journals publish many opinion, editorial, and journalistic pieces, and these could shape how science and evidence are perceived and what policies are adopted. These pieces are typically published quickly, often with little or no external review. Sometimes they may reflect overt advocacy that may increase the danger of bias and polarization of the scientific community.^5^ As more journals move towards publishing more opinion and advocacy, ethical guidelines are warranted.^6^

During the COVID-19 pandemic, science-based advocacy was common.^7^ While some argued for milder mitigation with restricted measures focused primarily on those at highest risk (e.g., the Great Barrington Declaration (GBD)^8,9^), others argued for mass suppression of the virus (e.g., the John Snow memorandum (JSM)^10^) or for elimination using aggressive lockdown measures, intense testing and contact tracing, social distancing, masking, and air monitoring and air cleaning interventions (“zeroCovid”).^11,12^ Understanding the presence of this advocacy in leading medical journals, given the historical importance of the issues involved, may help inform development of better guidelines for science-based advocacy in medical journals.

Here, we aimed to quantify the potential COVID-19 advocacy bias in the *BMJ*. *BMJ* is a leading journal with tremendous influence worldwide, arguably the premier journal championing evidence-based medicine with rigorous methods and protection from bias and conflicts of interest. We evaluated the share of advocates, editors, journalists, and independent scientists among the most prolific authors of COVID-19-related work in the *BMJ*; and assessed how *BMJ* published items authored by publicly declared advocates of aggressive measures (those who were leading members of both the Independent Scientific Advisory Group for Emergencies (indieSAGE)^13,14^ and the Vaccines-Plus initiative)^15^ relative to other scientist groups.

## METHODS

### Design

This is an exploratory meta-research analysis, and thus no protocol was pre-registered. We explored two research questions: 1) whether some advocacy was enriched in *BMJ* relative to the most prolific authors of COVID-19-related papers with UK addresses in the general literature (enrichment analysis); and 2) how strongly the dominant advocate group outperformed other groups of scientists in numbers of *BMJ* publications(controlled advocacy bias analysis) before (2016-2019) and during (2020-2023) the pandemic. We followed the STROBE guidelines in reporting the controlled comparison.

### Advocacy groups of interest

We focused on advocacy groups with clear, visible presence and public listing of key members. In the bibliometric analysis of prolific authors, we studied eight main advocacy groups: 1) indieSAGE members/key advisors (UK-based)^14^; 2) World Health Network (WHN) advocates defined as co-signatories on the Lancet WHN letter^16^ (this group is led from the US but also advocated elimination and its advocacy letter from October 2021 features many indieSAGE advocates); 3) advocates on the Vaccines-Plus letter, which contains UK and non-UK advocates but was initiated by UK indieSAGE advocates^15^; 4) JSM co-signatories on the original *Lancet* paper (contains non-UK signatories but was initiated by scientists affiliated with ZeroCovid advocacy); 5) GBD; 6) UK-led CollateralGlobal; 7) UK-based UsForThem; 8) UK-led Health Advisory & Recovery Team (HART). Groups 1-4 advocated for more aggressive policies whereas groups 5-8 advocated for more restricted policies. Details on the eight groups are summarized in Supplementary Text in the Supporting Information file.

### Data extraction

Data were extracted by two researchers independently (for the SCOPUS data: JPAI and IAC; for the *BMJ* data: KPK and JPAI) and discrepancies were discussed with a third researcher (KPK or IAC, respectively). A predesigned data extraction was developed that included use of the *BMJ* web site’s advanced search method with manual inspection of all collected documents to reduce misclassification. Information collected included the *BMJ* ID of the publication, the document type, the authors, and the publication year. Details of specific data protocols and extracted data are described below.

### Initial bibliometric analysis: most prolific COVID-19 authors

We searched Scopus for items published in the *BMJ* until April 13, 2024 on COVID-19 using the search string “COVID-19 OR SARS-CoV-2” in all fields. The most prolific authors were checked for being at any time members (indieSAGE webpage, HART group, UsForThemUK) or co-signatories (main authors the advocacy letters) of the eight initiatives listed above; editors or journalists; or, if none of these, other scientists. Members of official organisations like the WHO and the Royal Society of Medicine and patient interest groups not clearly aligned with a pandemic policy were included in the “other scientists” group. The analysis focused on those with 6 or more COVID-19-related publications in the *BMJ*, with special emphasis also on those with >10 such publications.

We also evaluated in Scopus the 100 most prolific scientists (excluding journalists) on COVID-19 for publications (in any scientific publication venue) with an address from the UK to examine the relative representation of advocates among them.

### Bibliometric analysis: controlled comparisons

Given the strong presence of indieSAGE and aligned advocacy groups in the retrieved documents, we performed additional analysis to investigate if the frequency of authorships and publications were biased. In order to ensure that we analyzed clear rather than short-term or low-commitment advocacy, our primary group of interest included the 16 scientists who were both members or key advisors of indieSAGE and co-authors of the Vaccines-Plus advocacy letter. We specifically evaluated whether indieSAGE/Vaccines-Plus advocates published more papers in *BMJ* during the pandemic years 2020-2023 as compared with the pre-pandemic years 2016-2019, and compared with other control groups of authors.

We considered four control groups, aiming to have exactly n=16 authors in each for balance against the indieSAGE/Vaccines-Plus group: The first control group included the 16 members of SAGE who attended at least four of the first 9 meetings of SAGE in early 2020.^17^ This comparison contrasts official government advisers versus self-organized advocates.

The second control group included the 16 scientists with a UK affiliation who were the most-highly cited according to a database of composite citation indicators^18^ and whose primary field in that database was considered to be relevant to *BMJ* (General & Internal Medicine; Epidemiology; Neurology & Neurosurgery; Nutrition & Dietetics; Respiratory System; Substance Abuse; Cardiovascular System & Hematology; Developmental & Child Psychology; Psychology; Statistics; Psychiatry; Immunology). We used the most updated citation database that focuses on the citation impact in a single most recent calendar year (2022) rather than whole career-long impact, so as to capture contemporary impact. This comparison contrasts advocates against the most extremely highly-cited scientists.

The third control group included the 16 UK-based scientists who are listed by name in the GBD website.^8^ This comparison contrasts two opposing advocacy groups with anti-diametric views.

The fourth control group included the 16 UK-based scientists who published the highest number of COVID-19 papers overall across all journals indexed in Scopus (excluding any indieSAGE members and current or previous editors such as Richard Horton from Lancet and Richard Smith and Fiona Godlee from *BMJ*). This comparison contrasted the advocates against a group with maximal interest in publishing COVID-19-related work.

For each author in the indieSAGE/Vaccines-Plus group and in each of the control groups, we examined whether they were among the top-2% most-cited authors in their field according to the composite citation indicator for single year impact in 2022^18^ and in 2019.^19^ We then counted the number of publications they had authored in *BMJ* each year between 2016 and 2023, using the advanced search method of the *BMJ* web site on author name (and affiliation, in cases of doubt), counting all document types. Membership of consortia also counted as authorship. The indieSAGE letter to *BMJ* was only counted to the main author, although other indieSAGE advocates cosigned it, given that it was merely a response to another paper. One letter listing indieSAGE as an author by itself was also not included. Given the large number of total counts, these two choices do not affect any conclusions of our work.

The search was done both for all document types without restriction, and restricted to COVID-19-related pieces (defined by presence of “COVID”, “SARS”, or “Pandemic” in the text, abstract, or title (using option: any word)). We also examined data for 2024 (up to April 20, 2024) to explore potentially differential evolution of publication patterns in the post-pandemic era. Publications were classified in four major groups: Original Research (Research), Review and Methods (Review, Research Methods and Reporting, Practice), Analysis (lengthy opinion pieces that may include also some data analyses) and Short Opinion (all other identified items: Views and reviews, Editorial, Opinion, Feature, Letter, Observation). Obituaries were not included in the analysis.

### Statistical analyses

We present descriptive statistics and avoid statistical testing of hypotheses given the exploratory nature of the evaluation.

## RESULTS

### Bibliometric analysis: most prolific COVID-19 authors

*BMJ* published 4,075 COVID-19-related items by April 13, 2024. 122 authors published more than 5 (up to 330 in the case of a journalist) COVID-19-related items each in the *BMJ* (**Table S2**). They included 18 advocates of aggressive policies (indieSAGE n=9, including a *BMJ* freelance journalist), WHN n=5, Vaccines-Plus n=12 (including the same *BMJ* freelance journalist), JSM main authors n=11 (20 if including low-level JSM co-signatories), with substantial overlap. The other prolific authors were 53 editors or journalists, and 51 other scientists not identified as associated with any advocacy. The 18 advocates of aggressive policies published 231 COVID-19-related papers in *BMJ*. An advocate who was a member of indieSAGE, left the organization in September 2020 and later joined CG that advocated for restricted measures. Since this advocate published 3 papers in 2020 before indieSAGE was formed, 4 while a member, and 6 after indieSAGE membership and the CG involvement started only in 2024, we classified this advocate as belonging to indieSAGE to avoid double counting. The prolific *BMJ* editors and journalists published about half of all *BMJ* COVID-19-related papers (n=564 and n=1355, respectively). Among 41 authors publishing >10 items in *BMJ*, 8 were advocates of aggressive policies, zero of restricted measures, 7 were editors, 23 journalists, and 3 were non-advocate scientists.

Conversely, among the 100 most prolific authors of COVID-19-related papers with a UK address, there were only 3 advocates of aggressive measures, 2 BMJ editors, 16 editors of other journals, and 79 other scientists (**Table S3**). If analysis was restricted to publications retrieved with COVID-19 OR SARS-CoV-2 in “Article title, Abstract, Keywords” rather than “All fields” in Scopus, the aggressive advocacy bias was similarly very large (**Table S2**). **Figure 1** summarizes these results on the representation of the studied advocates, editors, and journalists in *BMJ* versus the overall UK-based literature.

**Figure 1.**
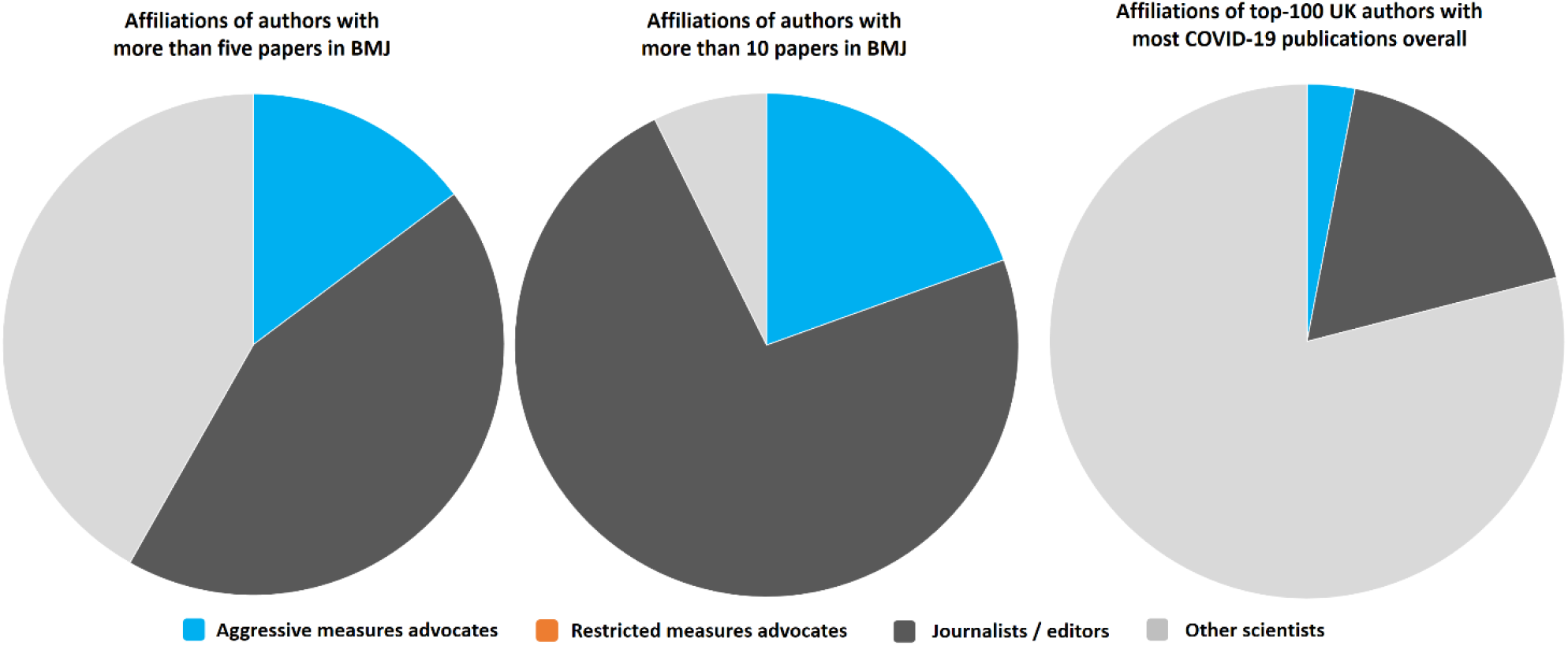
Most-prolific authors. Left: 122 authors with more than 5 *BMJ* COVID-19 related publications. Middle: 41 authors with more than 10 *BMJ* COVID-19-related publications. Right: 100 authors with 74 or more (up to 253) COVID-19-related publications with any UK address published in any venue (journalists excluded). One indieSAGE member subsequently advocated restricted measures (CG) in 2024; this special case was classified as aggressive measures advocacy, as best reflecting the *BMJ* publications of the study period.

### Bibliometric analysis: controlled comparisons

To understand whether the enrichment seen in Figure 1 also translated into an actual publishing bias, we report below a controlled publication analysis using five comparison groups to account for various types of confounding. All five groups of n=16 included excellent, high-impact scientists, as testified by the high proportion who were in the top-2% based on composite citation indicator data for their most recent year impact. For 2019 citation data, 9/16 in the indieSAGE/Vaccines-Plus group, 10/16 of the SAGE group, 8/16 of the GBD UK group, 16/16 of the UK most highly-cited group, and 12/16 in the group of UK scientists who published most COVID-19-related publications overall belonged to the top-2% most-cited scientists. In 2022 citation data, respective figures were 12/16, 10/16, 8/16, 16/16, and 16/16. (**Supplementary Table S4**).

**Table 1** shows the total number of authorships in the *BMJ* for the five groups. In the pre-pandemic period, the group of the most-cited scientists had the strongest presence in Research articles, while scientists who subsequently became indieSAGE/Vaccines-Plus advocates had quite strong presence in writing opinion pieces and scientists who subsequently became GBD advocates had practically no presence in the *BMJ*.

**Table 1.** Number of *BMJ* authorship appearances for indieSAGE/Vaccines-Plus advocates and four control groups, each with n=16 authors.

|  | **Total** | **Research** | **Opinion** | **Review/Methods** | **Analysis** |
| --- | --- | --- | --- | --- | --- |
| **2016-2019 (Pre-pandemic)** |  |  |  |  |  |
| IndieSAGE/Vaccines-Plus | 88 | 3 | 74 | 0 | 11 |
| SAGE | 4 | 1 | 2 | 0 | 1 |
| GBD UK | 0 | 0 | 0 | 0 | 0 |
| UK most highly cited | 32 | 20 | 7 | 3 | 2 |
| UK most COVID-19 papers | 9 | 3 | 5 | 1 | 0 |
| **2020-2023 (Pandemic)** |  |  |  |  |  |
| IndieSAGE/Vaccines-Plus | 385 | 18 | 330 | 7 | 30 |
| SAGE | 24 | 17 | 4 | 0 | 3 |
| GBD UK | 6 | 4 | 2 | 0 | 0 |
| UK most highly-cited | 24 | 6 | 14 | 3 | 1 |
| UK most COVID-19 papers | 36 | 25 | 8 | 2 | 1 |
| **2024 (Post-pandemic)** |  |  |  |  |  |
| IndieSAGE/Vaccines-Plus | 16 | 0 | 16 | 0 | 0 |
| SAGE | 0 | 0 | 0 | 0 | 0 |
| GBD UK | 0 | 0 | 0 | 0 | 0 |
| UK most highly-cited | 4 | 0 | 3 | 0 | 1 |
| UK most COVID-19 papers | 1 | 1 | 0 | 0 | 0 |

During pandemic years, indieSAGE/Vaccines-Plus advocates massively outperformed in *BMJ* presence all four control groups: 16.0-fold compared with leading SAGE members, 64.2-fold compared with the GBD advocates, 16.0-fold compared with the most-cited group, and 10.7-fold compared with the most prolific on COVID-19 group. The dominance of indieSAGE/Vaccines Plus advocates was most overwhelming in the Short Opinion group, where they outperformed the four control groups by 82.5-fold, 165-fold, 23.6-fold, and 41.3-fold, respectively; and in Analysis articles (10-30 fold, depending on comparison group). In the post-pandemic year, indieSAGE/Vaccines Plus authors seem to still publish many opinion pieces, while GBD authors have published nothing in the *BMJ* and the other three groups also had limited presence.

Although a minority of publications were coauthored by several advocates, when reducing the analysis to unique publications, a very similar picture was evident (**Table S5**). Comparing the pre-pandemic and pandemic years, although two indieSAGE/Vaccines-Plus associates already published many opinions in *BMJ* before 2020, the bias was massively enhanced during pandemic years (Figure 2).

**Figure 2.**
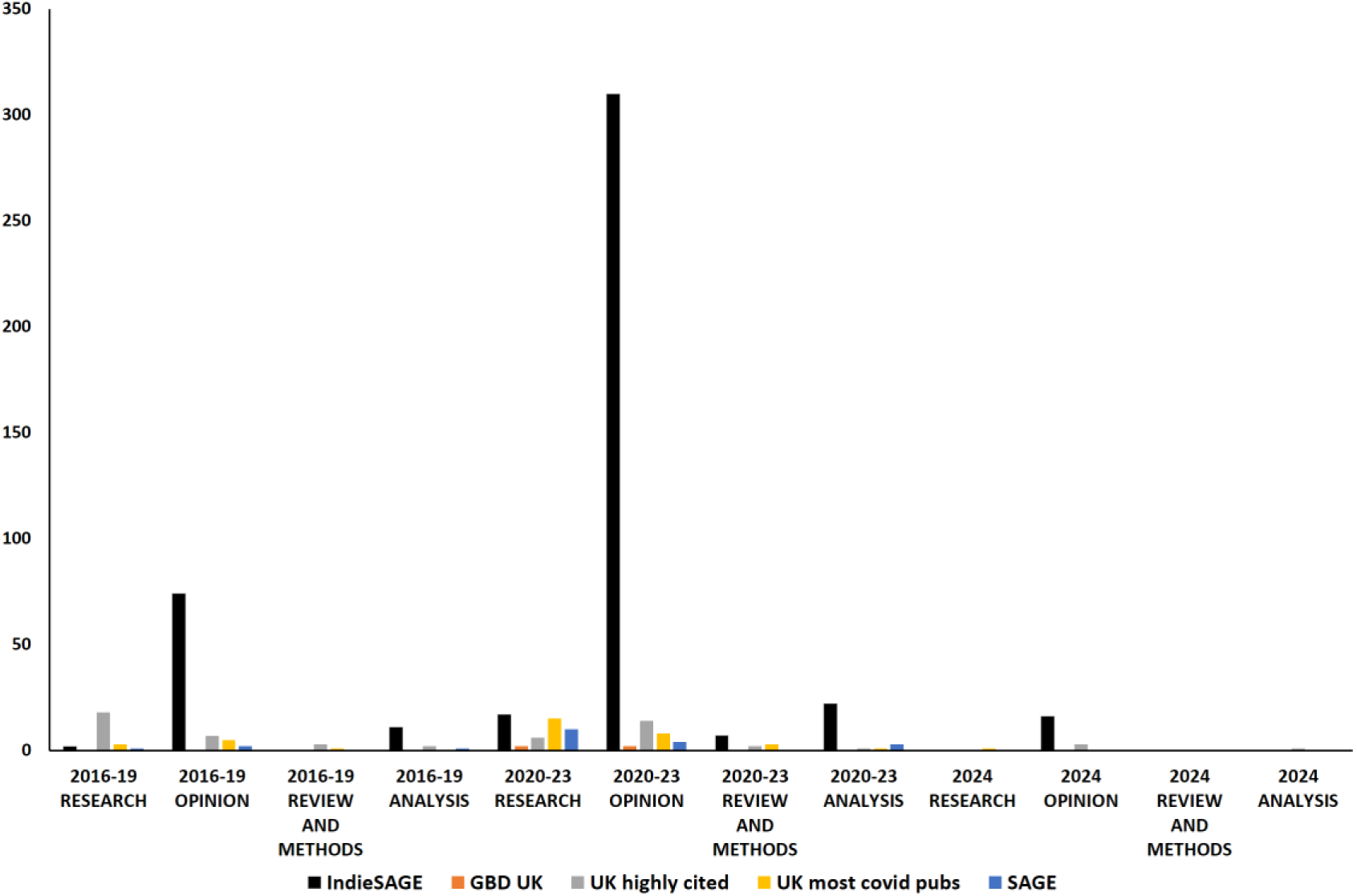
Historic development of unique number of *BMJ* publications featuring any member of the five studied groups of n=16 authors each (all publications with no content restriction)

A total 338 of 475 authorships (72%) during the pandemic years 2020-2023 of the members of the 5 groups were on COVID-19-specific publications. The extreme dominance of indieSAGE/Vaccines-Plus remained similar when analysis for 2020-2023 was limited to COVID-19-specific publications (**Table 2**).

**Table 2.** Number of *BMJ* authorship appearances for indieSAGE/Vaccines-Plus advocates and four control groups, each with n=16 authors, limited to published items that are COVID-19-specific.

|  | **Total** | **Research** | **Opinion** | **Review/Methods** | **Analysis** |
| --- | --- | --- | --- | --- | --- |
| **2020-2023 (Pandemic)** |  |  |  |  |  |
| IndieSAGE/Vaccines Plus | 272 | 17 | 221 | 7 | 27 |
| SAGE | 21 | 15 | 3 | 0 | 3 |
| GBD UK | 6 | 4 | 2 | 0 | 0 |
| UK most highly-cited | 11 | 1 | 9 | 0 | 1 |
| UK most COVID-19 papers | 28 | 20 | 5 | 2 | 1 |

The relative presence in the *BMJ* of the different indieSAGE/Vaccines-Plus members varied substantially. During 2020-2023, one member who also served as freelance journalist published 180 opinions and views (97 of them COVID-19-related) in the *BMJ*. Even without this author, the bias remained massive (**Tables S6, S7**). Among other indieSAGE/Vaccines-Plus members, one published in the same period 81 papers (69 COVID-19-related), and another published 29 (28 COVID-19-related), while the others had less prolific contributions.

## DISCUSSION

Our analysis suggests that *BMJ* massively published advocate authors championing zeroCOVID policies and later, other indieSAGE-led aggressive approaches to COVID-19 during the pandemic. Leading members of SAGE, highly-cited UK scientists and the most prolific researchers on COVID-19 across the entire scientific literature had very limited *BMJ* presence compared with the preferred advocates. Advocates of restricted, focused measures have been almost extinct from *BMJ* pages. *BMJ* editors, staff and apparently advocate contributors developed a massive literature, comprised mostly of opinion pieces that in general (as acknowledged by the *BMJ*) underwent no external review in the *BMJ*. The degree of apparent favoritism exhibited by what is considered to be the premier venue of evidence-based medicine is very concerning and invites further scrutiny.

Scientific journals have a responsibility to be balanced, objective, and factual, giving that endorsement of specific ideological or political positions may distort evidence and lead to polarization of the scientific community and loss of trust.^20^ The intense advocacy by indieSAGE in *BMJ* was accompanied by UK media publishing many views by indieSAGE, with almost 200 being available on IndieSAGE’s own web page, and these views were sometimes confused with the official SAGE in British media.^21,22^ This confusion led to exposure of the British population to zeroCOVID advocacy without appropriately recognizing it as such. Given the worldwide influence of *BMJ*, the impact of this distortion probably had global consequences.

Advocacy may be associated also with hostility towards other scientists, both on social media and in *BMJ*, promoting obsessive forms of criticism.^23^ In *BMJ*, SAGE modeling was held co-responsible for tens of thousands of deaths.^24^ On Twitter/X, UK scientists were also criticized intensely (**Table S8**). In a letter in *BMJ Evidence Based Medicine*,^25^ three WHN advocates called a paper on long COVID^26^ a “Trojan horse” and accused the authors of ideological biases, while themselves declaring no association with WHN or indieSAGE. A paper on “misinformation” in *BMJ*^27^ by advocates studied here that criticized some other groups also lacked these declarations.

Some limitations of our work should be discussed. First, we only evaluated one major journal, and similar assessments in other leading journals seem warranted as part of a more general post-pandemic meta-science evaluation. Discussion of journal-led science-based advocacy is needed. Leading journals with large, influential magazine sections, like the *BMJ*, are particularly important to study because they have major impact and can publish many opinion papers very quickly, while peer-reviewed research is far slower. *BMJ*, *Lancet*, *Nature*, and *Science* have many editors and journalists who may publish hundreds of items in their pages, typically without external peer-review and disclosure of conflicts of interest.^28^

The specific bias we observed here may also have occurred elsewhere. For example, the author who published most COVID-19-related publications in *BMJ* was also the most prolific academic in COVID-19-related publications in the *Lancet* (n=36 published items); and the second most-prolific in *BMJ* was also prolific in the *Lancet* (n=10 published items). Both were indieSAGE/Vaccines-Plus advocates, with most of their documents being opinion pieces. In the case of the *Lancet*, the highest number of COVID-19-related publications were anonymous editorials (n=65) or items authored by the editor-in-chief (n=50). The editor-in-chief himself also advertised “noCOVID” aggressive mitigation advocacy.^29^

Second, as we do not have submission statistics available, our study only informs on final published documents, but the large majority of submissions to *BMJ* are rejected. Editors and advocates may shape what gets published through the editorial and peer-reviewing process, and authors with views not congruent with zeroCOVID advocacy may even have stopped submitting to *BMJ* after seeing the overt bias that we describe here, or after receiving disparaging feedback. We encourage the *BMJ* to release information that could illuminate this, including how many papers the scientists analysed here were invited to review. In support of this concern, we preliminarily explored the available reviewer names of the 64 COVID-19-related externally reviewed *BMJ* Research, Analysis, and Review papers that were published by the 80 members of the 5 analysed groups of authors (17 Research and 22 Analysis for the indieSAGE/vaccines-Plus group and 37 by the other groups combined, which reduced to 25 unique documents after removing duplicates due to group authorship overlap). 9 of the 64 unique documents had been reviewed by at least 1 advocate of aggressive measures and 0 by advocates of restricted measures; of these 9 cases, 7 were papers by indieSAGE, i.e. advocates reviewed papers by other advocates belonging to the same advocacy group. For example, Greenhalgh reviewed Haque (BMJ 2021; 372: n693) and Scally and Kvalsvig reviewed McKee (BMJ 2021; 372: n208; BMJ 2022; 378: e069558). Two of the Analyses written by indieSAGE on masks^30^ and COVID-19 misinformation^27^ had their peer-review hidden against standard journal policy. These sparse data suggest strong advocacy collusion but they need to be augmented by reviewer information on all COVID-19 papers submitted to the *BMJ*, including rejected submissions. Given that space in a competitive journal is very limited, the many advocate Analysis papers in *BMJ* and the editorial commissioning of many indieSAGE opinions^31^ may have led to correspondingly less favorable reviewing experiences for other scientists.

Third, the comparisons made here have various confounding effects. Dedicated advocates are by their very call to advocacy more likely to publish opinions. Our analysis has attempted to account for this confounding by comparing to diverse groups of other authors, including leading GBD scientists, who are also expected to have a call to advocacy. One of the indieSAGE advocates was a *BMJ* freelance journalist publishing 180 opinions and views during 2020-23, many proposing wide-reaching public health policies with little or no evidence. Although this by itself raises questions and contributes to the massive bias observed here, the bias remains massive if this journalist is removed from the analysis (**Tables S6-S7**). One could argue that SAGE as a comparison group has less inclination to publish pandemic opinion pieces by their role as official advisors; we controlled for this by comparing also to other highly-cited UK scientists and non-advisor scientists with a clear research interest in COVID-19. The fact that the bias was massive regardless of what control group we used shows that all these potential confounders have limited effect compared to the total observed bias signal.

Fourth, we did not aim to appraise whether the claims and policy proposals of the advocates were correct or wrong, or if the methods used were worse than any other science-related advocacy in circulation. However, the status of elimination policies as a minority position in the scientific community^32^ and the eventual infeasibility of zeroCOVID^33^ contrast strongly to the special preference that advocates of this position had in *BMJ*. This suggests that the bias was not only misplaced in quantitative terms but also misplaced or even devastatingly wrong in qualitative terms.

Fifth, we only examined advocates based on highly visible, uncontestable advocacy groups, but there are several other organizations, movements, and initiatives that advocated and lobbied during the pandemic, often without having publicly listed memberships. Therefore, advocacy infiltration of the literature may be more prominent than what we observed.

Some suggestions for the future can be made based on our analysis. First, editors should consider placing a cap on how often they can host opinion pieces of any particular scientist (or even their own views) in a given calendar year. Editorial nepotism has been described to be a widespread problem.^34^ Massive publication of non-evidence-based opinions by editors or favored authors could distort consensus on available evidence in some critical circumstances. Original research articles rarely affect public policy by themselves, while opinions in major journals set the tone for far-reaching policy choices. Second, journals where massive advocacy bias and other forms of favoritism are demonstrated may wish to establish independent auditing to examine whether collusion affected editorial practices. Third, readers and the general public should be sensitized to these problems so as to avoid being misled in the future. This requires new empirical studies and clarified principles regulating science communication. Finally, journals may need to ensure space for debate articles where different views are juxtaposed, each supported by evidence, in the best interest of science and evidence-based policy-making.

## Data sharing statement

All data used in this study are publicly available at *BMJ* or can be obtained via SCOPUS. We provide relevant raw data in Supplementary information. If any data or calculations remain unclear, readers are warmly welcome to contact the authors.

## Transparency declaration

The lead author (JPAI) affirms that the manuscript is an honest, accurate, and transparent account of the study being reported; that no important aspects of the study have been omitted. All data and calculations are available upon request.

## Ethics approval

The work described did not require ethics approval as it is a bibliometric analysis.

## Data Availability

All data used in this study are publicly available at BMJ or can be obtained via SCOPUS. We provide relevant raw data in Supplementary information. If any data or calculations remain unclear, readers are warmly welcome to contact the authors.

## SUPPORTING INFORMATION

### Supplementary methods

Of the four advocacy groups favoring aggressive measures, IndieSAGE^34^ was formed in May 2020 to provide an independent, critical counterpart to the Scientific Advisory Group for Emergencies (SAGE), the official UK governmental COVID-19 scientific advisory taskforce. IndieSAGE advocated COVID-19 elimination, i.e., a “zeroCOVID” strategy.^34^ Membership in indieSAGE changed over time (**Table S1**) and not all members were equally active in advocacy while some members also changed views and/or left indieSAGE. Like indieSAGE, the WHN, initated in 2020 and led by Yaneer Bar-Yam and Eric Ding, also advocated for elimination (“zero-covid”) strategies.^34^ Vaccines-Plus published a manifesto^34^ in *BMJ* in January 2022, when zeroCOVID was clearly no longer feasible, arguing for efforts to tightly control infections using “effective find, test, trace, isolate, and support” strategies, use of respirators (e.g. N95, P2/FFP2, KF94) in all indoor settings, and aiming for “a paradigm shift to ensure all public buildings are designed, built, adapted, and utilised to maximise clean air”, while also advocating (less controversial) global vaccine equity. Advocates in these groups had substantial overlap with the key authors of JSM in *Lancet*.^34^

Of the four advocacy groups favoring restricted measures, GBD^34^ was initiated by one UK and two US scientists, advocated for “focused” protection, i.e. giving more freedom to younger age groups, including school children. This group has many UK scientists in the list of publicly visible key signatories. CollateralGlobal was initiated by UK scientists and emphasizes increased focus on the adverse effects of pandemic mitigation policies. We used the list of signatories of a letter questioning the UK COVID inquiry.^34^ UsForThemUK emphasized normalcy of school kids during the pandemic but later advocated also a broader range of policies.^34^ Finally, HART was a (very) low-intervention pandemic advocacy group that strongly favored keeping children and schools out of the pandemic mitigation and also criticized the mass vaccination policies during the pandemic.^34^

Besides the analysis of main advocates, JSM provides also a list of 4,200 people who co-signed it besides the main author co-signatories.^34^ As a supplementary analysis, the presence of such additional co-signatories was assessed in the most-prolific COVID-19-related authors in the *BMJ* and in the most-prolific COVID-19-related authors in papers with UK address. GBD, that JSM opposed, did not provide in public full lists of secondary co-signatories by name. The long list of 4,200 JSM co-signatories included additionally 9 who were among the most prolific in the *BMJ* (**Table S2**) and additionally 7 who were among the most prolific in papers with a UK address (**Table S3**), indicating that, in stark contrast to the enrichment of the “high-level” advocacy discussed above, only a modest, arguably non-significant, *BMJ* enrichment in “low-level” advocacy was seen.

**Table S1.** Members of indieSAGE, and alleged period of membership.^a^.

| Member Period | Name |
| --- | --- |
| Dec 2021 – Oct 2023 | Altmann, Daniel |
| May 2020 – | Costello, Anthony |
| April 2022 – | Cruickshank, Sheena |
| May 2020 – May 2022 | Friston, Karl |
| April 2022 | Greenhalgh, Trisha |
| Jan 2022 – | Griffin, Stephen |
| April 2022 – May 2022 | Gurdasani, Deepti |
| May 2020 – | Haque, Zubaida |
| April 2022 – | Kane, Binita |
| Jan 2022 – | Katzourakis, Aris |
| May 2020 – May 2021 | Khunti, Kamlesh |
| May 2020 – August 2021 | King, David A. |
| Jan 2023 – | Lee, Lennard |
| May 2020 – | McKee, Martin |
| May 2020 – | Michie, Susan |
| June 2020 – Feb 2022 | Oni, Tolullah |
| May 2020 – | Pagel, Christina |
| May 2020 – Dec 2021; Apr 2022 – October 2022 | Pillay, Deenan |
| May 2020 | Pittard, Alison |
| May-September 2020 | Pollock, Allyson |
| June 2020 – | Reicher, Stephen |
| April 2022 – | Robertson, Duncan |
| August 2021 – | Salisbury, Helen |
| May 2020 – | Scally, Gabriel |
| October 2020- | Yates, Christian/Kit |

<sup>a</sup> Data taken from indieSAGE's web page (https://www.independentsage.org/who-are-independent-sage/)
https://web.archive.org/web/20240221090620/https://www.independentsage.org/who-are-independent-sage/

**Table S2.**
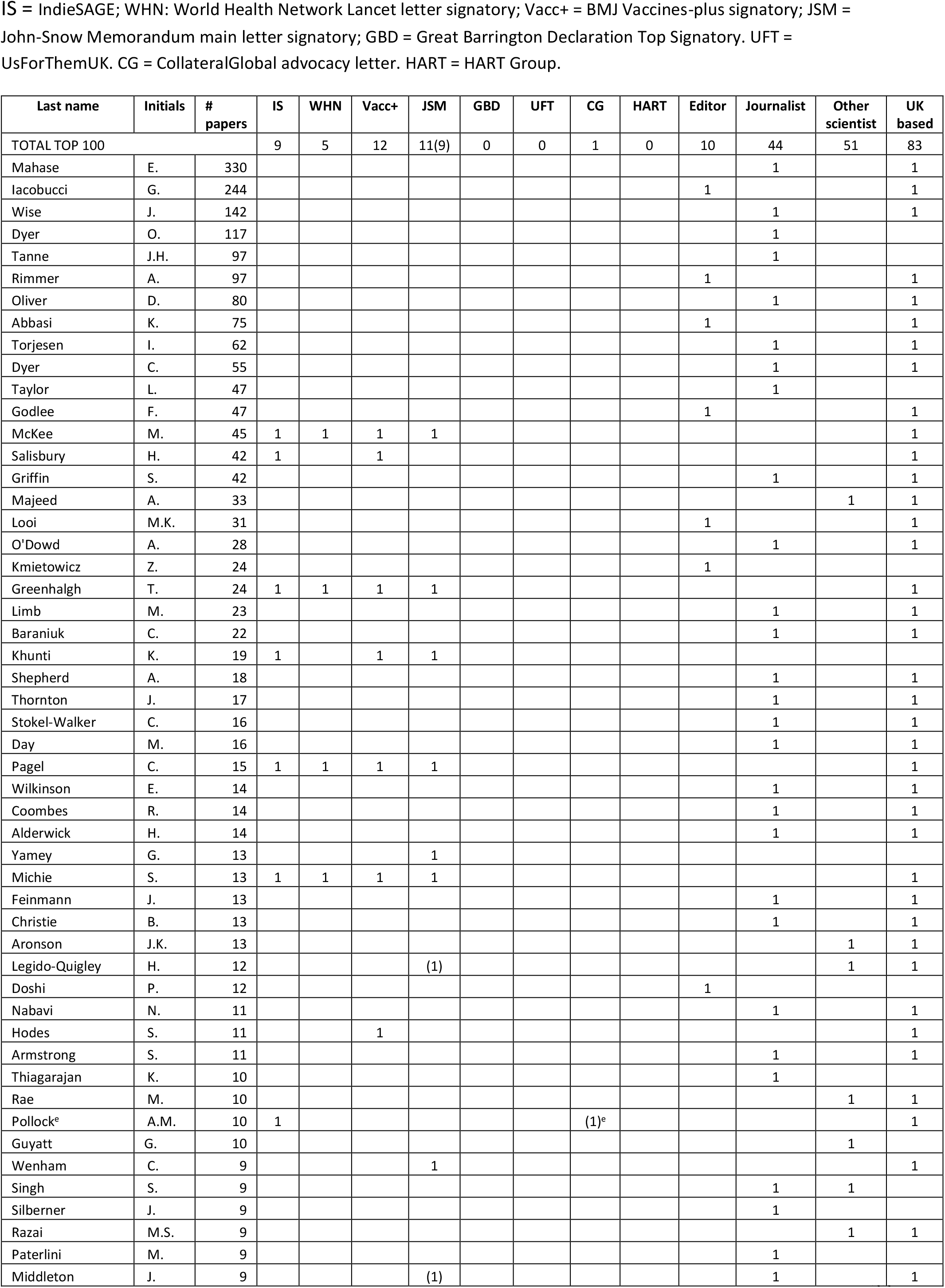
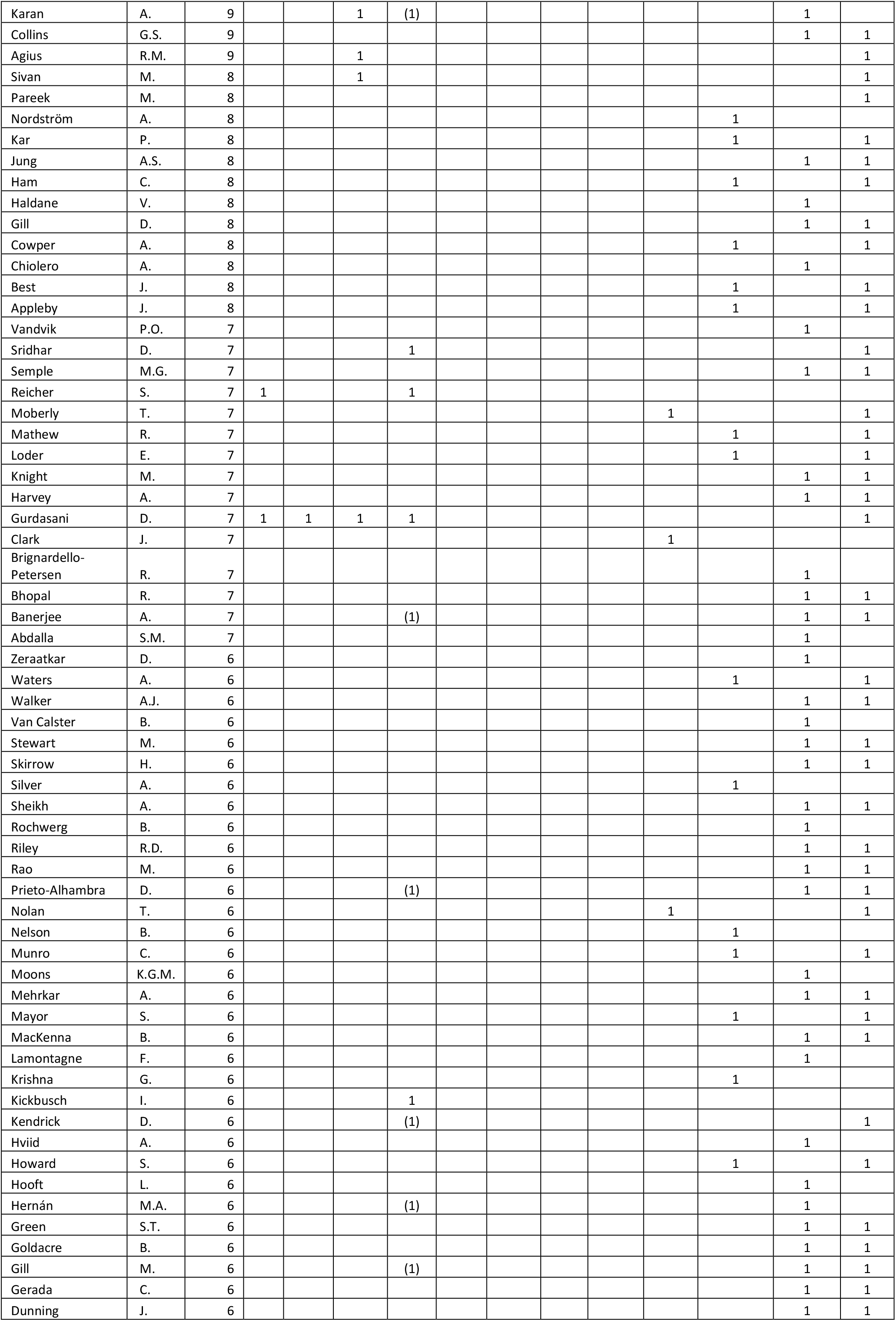
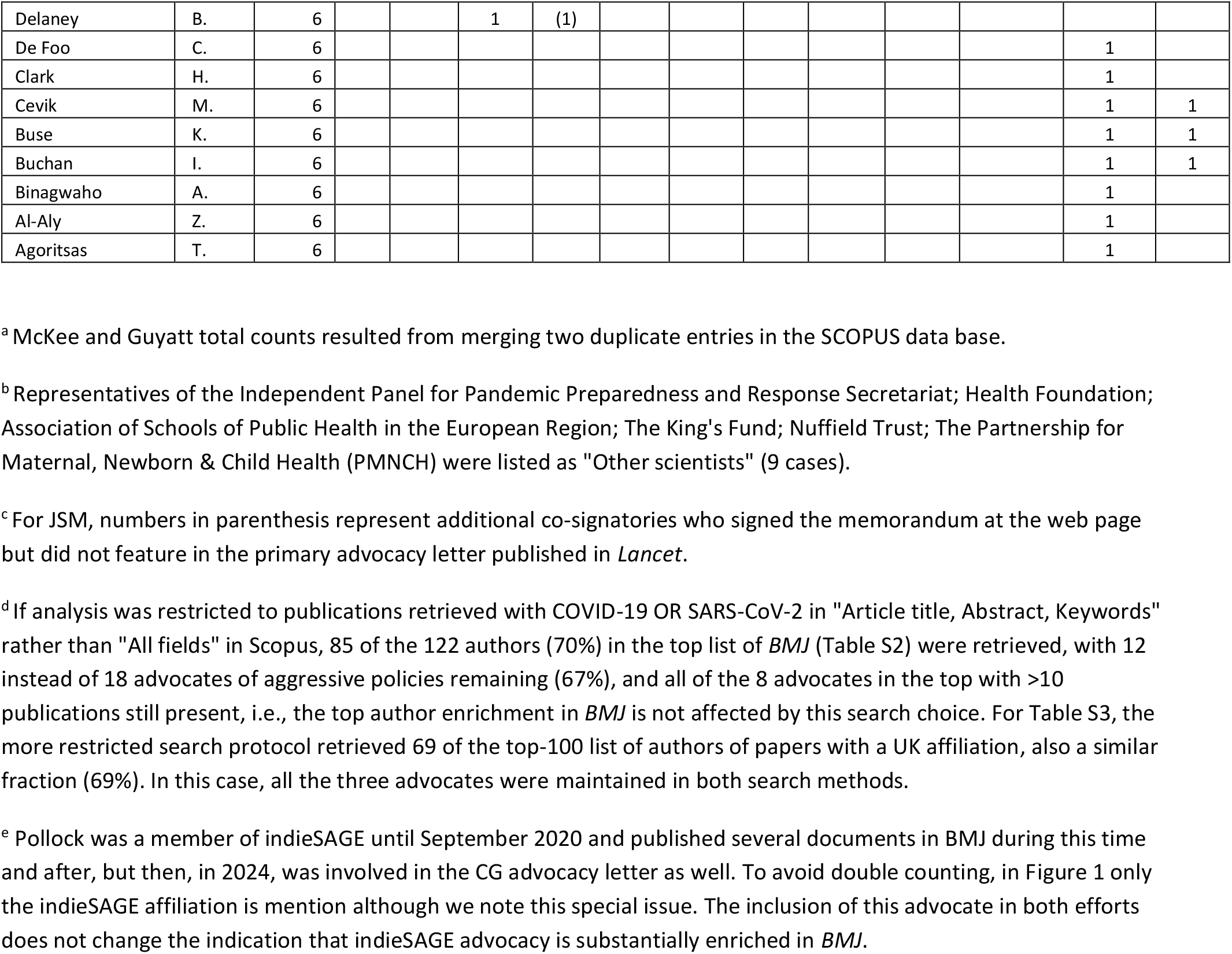
Affiliations of 122 authors publishing more than 5 papers in BMJ.^a,b,c,d,e^. IS = IndieSAGE; WHN: World Health Network Lancet letter signatory; Vacc+ = BMJ Vaccines-plus signatory; JSM = John-Snow Memorandum main letter signatory; GBD = Great Barrington Declaration Top Signatory. UFT = UsForThemUK. CG = CollateralGlobal advocacy letter. HART = HART Group.

**Table S3.** Affiliations of Top-100 UK authors publishing most on COVID-19.^a,b,c,d,e^. IS = IndieSAGE; WHN: World Health Network Lancet letter signatory; Vacc+ = BMJ Vaccines-plus signatory; JSM = John-Snow Memorandum main letter signatory; GBD = Great Barrington Declaration Top Signatory. UFT = UsForThemUK. CG = CollateralGlobal advocacy letter. HART = HART Group.

| Last name | Initials | # papers | IS | WHN | Vacc+ | JSM | GBD | UFT | CG | HART | Editor | Journalist | Other scientist |
| --- | --- | --- | --- | --- | --- | --- | --- | --- | --- | --- | --- | --- | --- |
| TOTAL TOP 100 |  |  | 3 | 2 | 3 | 3(7) | 0 | 0 | 0 | 0 | 18 | 0 | 79 |
| Khunti | K. | 253 | 1 |  | 1 | 1 |  |  |  |  |  |  |  |
| Griffiths | M.D. | 248 |  |  |  |  |  |  |  |  |  |  | 1 |
| Smith | L. | 178 |  |  |  |  |  |  |  |  |  |  | 1 |
| Hasan | S.S. | 173 |  |  |  |  |  |  |  |  |  |  | 1 |
| Zumla | A. | 170 |  |  |  |  |  |  |  |  |  |  | 1 |
| McKee | M. | 166 | 1 | 1 | 1 | 1 |  |  |  |  |  |  |  |
| Godlee<sup>b</sup> | F. | 160 |  |  |  |  |  |  |  |  | 1 |  |  |
| Horton<sup>b</sup> | R. | 157 |  |  |  | (1) |  |  |  |  | 1 |  |  |
| Kow | C.S. | 153 |  |  |  |  |  |  |  |  |  |  | 1 |
| Sheikh | A. | 216 |  |  |  |  |  |  |  |  |  |  | 1 |
| Koyanagi | A. | 152 |  |  |  |  |  |  |  |  |  |  | 1 |
| Laybourn-Langton<sup>b</sup> | L. | 146 |  |  |  |  |  |  |  |  | 1 |  |  |
| Smith<sup>b</sup> | R. | 145 |  |  |  |  |  |  |  |  | 1 |  |  |
| Norman<sup>b</sup> | I. | 143 |  |  |  |  |  |  |  |  | 1 |  |  |
| Atwoli<sup>b</sup> | L. | 142 |  |  |  |  |  |  |  |  | 1 |  |  |
| Rubin<sup>b</sup> | E.J. | 141 |  |  |  |  |  |  |  |  | 1 |  |  |
| Praities<sup>b</sup> | N. | 140 |  |  |  |  |  |  |  |  | 1 |  |  |
| Turale<sup>b</sup> | S. | 139 |  |  |  |  |  |  |  |  | 1 |  |  |
| Sahni<sup>b</sup> | P. | 139 |  |  |  |  |  |  |  |  | 1 |  |  |
| Vázquez<sup>b</sup> | D. | 138 |  |  |  |  |  |  |  |  | 1 |  |  |
| Patrick<sup>b</sup> | K. | 138 |  |  |  |  |  |  |  |  | 1 |  |  |
| Hancocks<sup>b</sup> | S. | 138 |  |  |  |  |  |  |  |  | 1 |  |  |
| Bosurgi<sup>b</sup> | R. | 138 |  |  |  |  |  |  |  |  | 1 |  |  |
| Zambon | M. | 133 |  |  |  |  |  |  |  |  |  |  | 1 |
| Godman | B. | 131 |  |  |  |  |  |  |  |  |  |  | 1 |
| Monteiro<sup>b</sup> | C.A. | 130 |  |  |  |  |  |  |  |  | 1 |  |  |
| Semple | M.G. | 129 |  |  |  |  |  |  |  |  |  |  | 1 |
| Lucero-Prisno | D.E. | 129 |  |  |  |  |  |  |  |  |  |  | 1 |
| Jit | M. | 128 |  |  |  | (1) |  |  |  |  |  |  | 1 |
| Wong | I.C.K. | 127 |  |  |  |  |  |  |  |  |  |  | 1 |
| Baqui<sup>b</sup> | A.H. | 124 |  |  |  |  |  |  |  |  | 1 |  |  |
| Dwivedi | Y.K. | 123 |  |  |  |  |  |  |  |  |  |  | 1 |
| Banerjee | A. | 122 |  |  |  | (1) |  |  |  |  |  |  | 1 |
| Ladhani | S.N. | 121 |  |  |  |  |  |  |  |  |  |  | 1 |
| Klenerman | P. | 119 |  |  |  |  |  |  |  |  |  |  | 1 |
| Katikireddi | S.V. | 119 |  |  |  |  |  |  |  |  |  |  | 1 |
| Eggo | R.M. | 117 |  |  |  |  |  |  |  |  |  |  | 1 |
| Benfield<sup>b</sup> | T. | 117 |  |  |  |  |  |  |  |  | 1 |  |  |
| Hopkins | C. | 113 |  |  |  |  |  |  |  |  |  |  | 1 |
| Lip | G.Y.H. | 112 |  |  |  |  |  |  |  |  |  |  | 1 |
| Pollard | A.J. | 111 |  |  |  |  |  |  |  |  |  |  | 1 |
| Fancourt | D. | 108 |  |  |  |  |  |  |  |  |  |  | 1 |
| Baillie | J.K. | 107 |  |  |  |  |  |  |  |  |  |  | 1 |
| Greenhalgh | T. | 106 | 1 | 1 | 1 | 1 |  |  |  |  |  |  |  |
| Donnelly | C.A. | 103 |  |  |  |  |  |  |  |  |  |  | 1 |
| Majeed | A. | 102 |  |  |  |  |  |  |  |  |  |  | 1 |
| Turtle | L. | 101 |  |  |  |  |  |  |  |  |  |  | 1 |
| Lambe | T. | 101 |  |  |  |  |  |  |  |  |  |  | 1 |
| Darzi | A. | 101 |  |  |  |  |  |  |  |  |  |  | 1 |
| Rambaut | A. | 99 |  |  |  | (1) |  |  |  |  |  |  | 1 |
| Pareek | M. | 99 |  |  |  |  |  |  |  |  |  |  | 1 |
| Shankar-Hari | M. | 97 |  |  |  |  |  |  |  |  |  |  | 1 |
| Chand | M. | 97 |  |  |  |  |  |  |  |  |  |  | 1 |
| Mahmud | M. | 96 |  |  |  |  |  |  |  |  |  |  | 1 |
| Camporota | L. | 96 |  |  |  |  |  |  |  |  |  |  | 1 |
| Lyons | R.A. | 95 |  |  |  |  |  |  |  |  |  |  | 1 |
| Robertson | C. | 94 |  |  |  |  |  |  |  |  |  |  | 1 |
| Lin | C.Y. | 94 |  |  |  |  |  |  |  |  |  |  | 1 |
| de Lusignan | S. | 93 |  |  |  |  |  |  |  |  |  |  | 1 |
| Akbari | A. | 92 |  |  |  |  |  |  |  |  |  |  | 1 |
| Thomson | E.C. | 91 |  |  |  |  |  |  |  |  |  |  | 1 |
| Jacob | L. | 91 |  |  |  |  |  |  |  |  |  |  | 1 |
| Funk | S. | 91 |  |  |  | (1) |  |  |  |  |  |  | 1 |
| Zhang | Y.D. | 90 |  |  |  |  |  |  |  |  |  |  | 1 |
| Deveci | M. | 90 |  |  |  |  |  |  |  |  |  |  | 1 |
| Solomon | T. | 89 |  |  |  |  |  |  |  |  |  |  | 1 |
| Schuller | B.W. | 88 |  |  |  |  |  |  |  |  |  |  | 1 |
| Openshaw | P.J.M. | 88 |  |  |  |  |  |  |  |  |  |  | 1 |
| Mentzer | A.J. | 88 |  |  |  |  |  |  |  |  |  |  | 1 |
| Kumar | A. | 88 |  |  |  |  |  |  |  |  |  |  | 1 |
| Sigfrid | L. | 86 |  |  |  |  |  |  |  |  |  |  | 1 |
| Ostermann | M. | 86 |  |  |  |  |  |  |  |  |  |  | 1 |
| Sullivan | R. | 85 |  |  |  |  |  |  |  |  |  |  | 1 |
| Summers | C. | 84 |  |  |  |  |  |  |  |  |  |  | 1 |
| Khalil | A. | 84 |  |  |  |  |  |  |  |  |  |  | 1 |
| Harrison | E.M. | 84 |  |  |  |  |  |  |  |  |  |  | 1 |
| Barclay | W.S. | 84 |  |  |  |  |  |  |  |  |  |  | 1 |
| Onyeaka | H. | 83 |  |  |  |  |  |  |  |  |  |  | 1 |
| Kotera | Y. | 83 |  |  |  |  |  |  |  |  |  |  | 1 |
| Yon | D.K. | 81 |  |  |  |  |  |  |  |  |  |  | 1 |
| Harky | A. | 81 |  |  |  |  |  |  |  |  |  |  | 1 |
| Szakmany | T. | 80 |  |  |  |  |  |  |  |  |  |  | 1 |
| Kucharski | A.J. | 80 |  |  |  |  |  |  |  |  |  |  | 1 |
| Aujayeb | A. | 80 |  |  |  |  |  |  |  |  |  |  | 1 |
| Dabrera | G. | 79 |  |  |  |  |  |  |  |  |  |  | 1 |
| Abbott | S. | 79 |  |  |  |  |  |  |  |  |  |  | 1 |
| Schultz | M.J. | 78 |  |  |  |  |  |  |  |  |  |  | 1 |
| Kraemer | M.U.G. | 78 |  |  |  |  |  |  |  |  |  |  | 1 |
| Gupta | L. | 78 |  |  |  |  |  |  |  |  |  |  | 1 |
| Dunning | J. | 78 |  |  |  |  |  |  |  |  |  |  | 1 |
| Barnes | E. | 78 |  |  |  |  |  |  |  |  |  |  | 1 |
| Olde Rikkert<sup>b</sup> | M.G.M. | 77 |  |  |  |  |  |  |  |  | 1 |  |  |
| Finn | A. | 77 |  |  |  |  |  |  |  |  |  |  | 1 |
| Tang | J.W. | 76 |  |  |  |  |  |  |  |  |  |  | 1 |
| Shin | J.I. | 76 |  |  |  |  |  |  |  |  |  |  | 1 |
| Pybus | O.G. | 76 |  |  |  |  |  |  |  |  |  |  | 1 |
| Prieto-Alhambra | D. | 75 |  |  |  | (1) |  |  |  |  |  |  | 1 |
| Hopkins | S. | 75 |  |  |  |  |  |  |  |  |  |  | 1 |
| Bhatt | S. | 75 |  |  |  | (1) |  |  |  |  |  |  | 1 |
| Noursadeghi | M. | 74 |  |  |  |  |  |  |  |  |  |  | 1 |

<sup>a</sup> A. Sheikh and A. Koyanagi total counts resulted from merging two duplicate entries in the SCOPUS data base.
<sup>b</sup> Many of the publications by these editors were the same climate policy position papers that appeared in multiple journals.
<sup>c</sup> For JSM, numbers in parenthesis represent additional co-signatories who signed the memorandum at the web page but did not feature in the primary advocacy letter published in *Lancet*.
<sup>d</sup> We note that an indieSAGE advocate (Michie) is number 101 (the next entry) on this list, but even with this advocate included, the enrichment in *BMJ* (Table S2) over the general literature (Table S3) is still very substantial.
<sup>e</sup> For Table S3, the more restricted search protocol retrieved 69 of the top-100 list of authors of papers with a UK affiliation (69%). In this case, all the three advocates listed were maintained in both search methods.

**Table S4.**
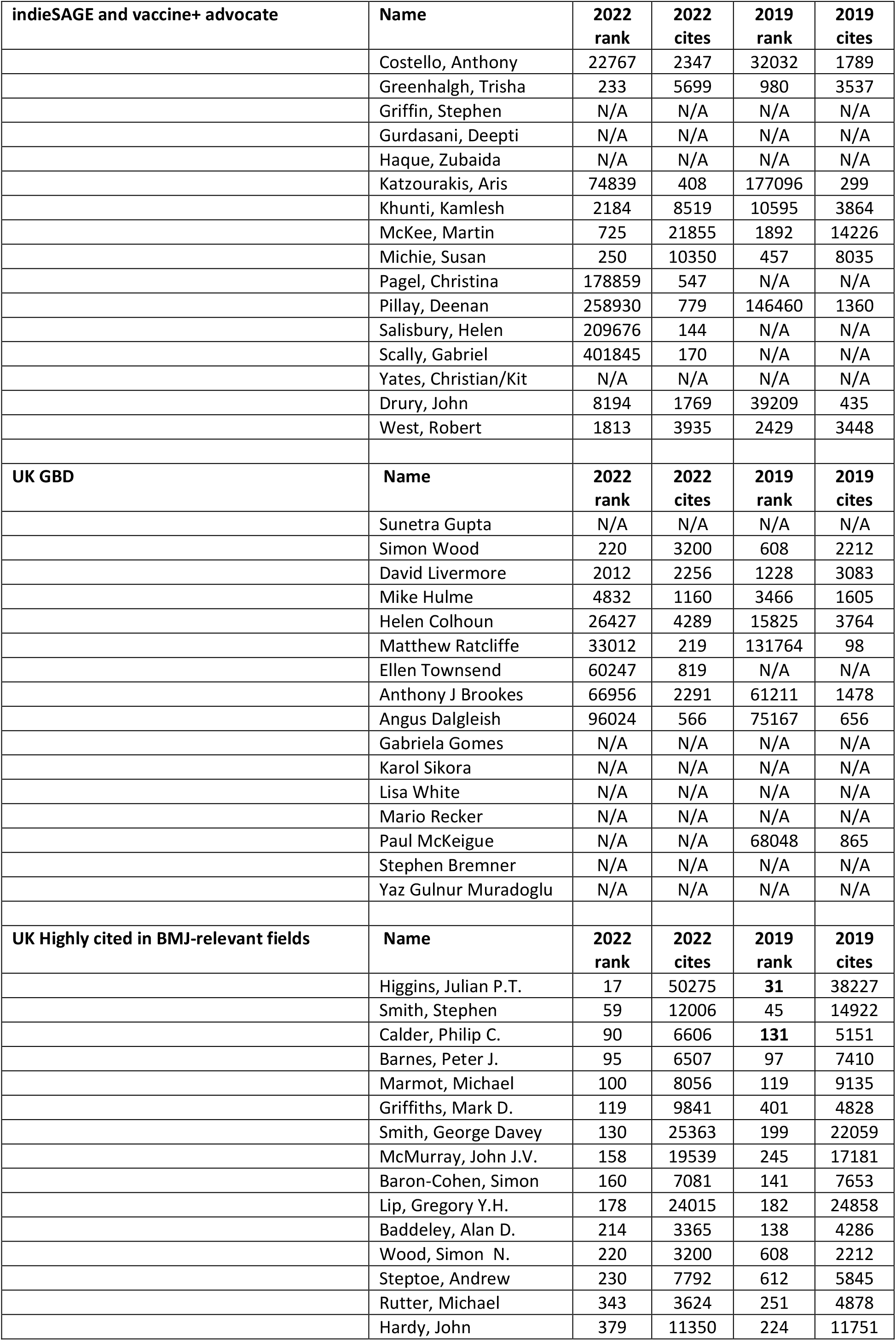
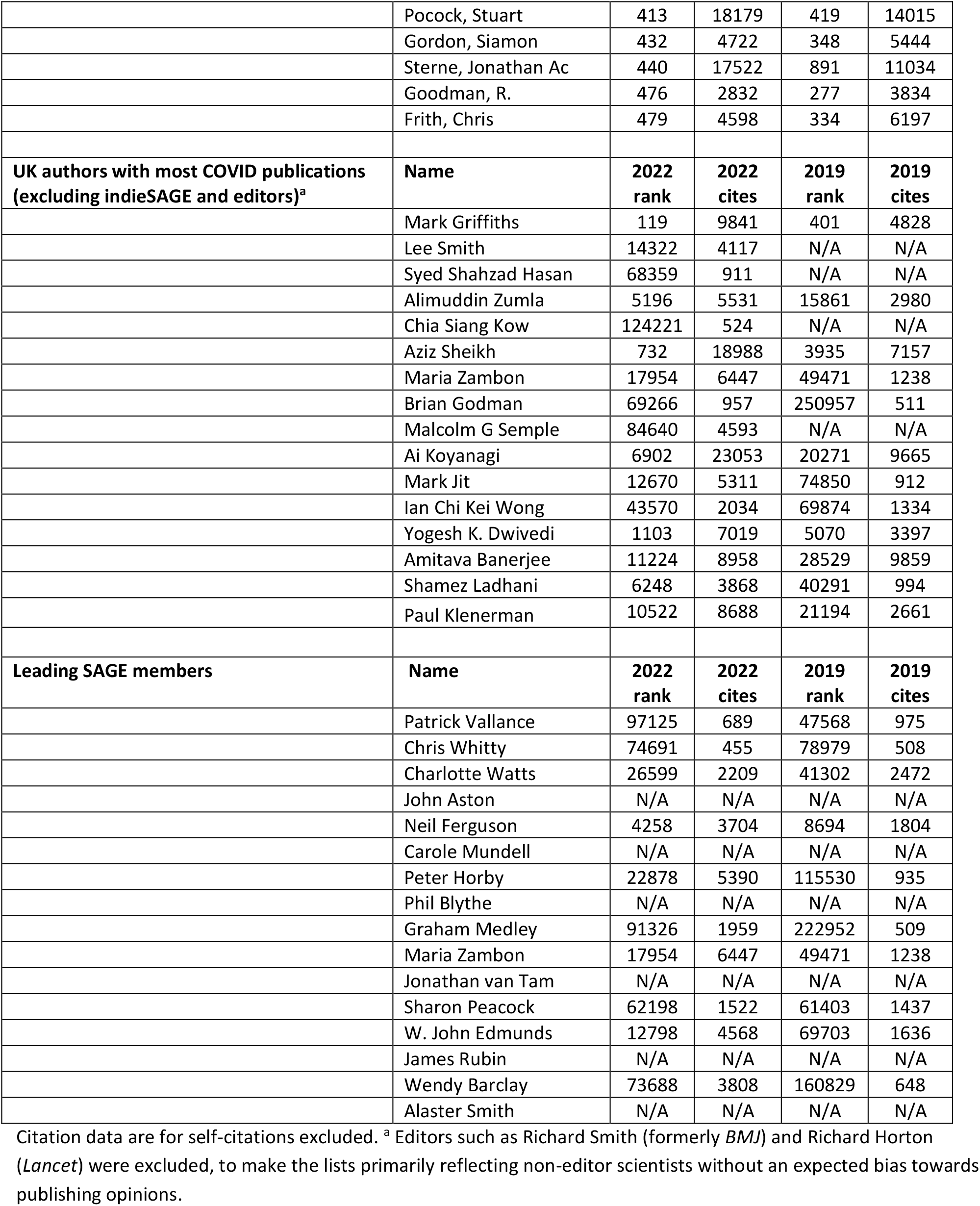
Comparison groups of 16 used for controlled comparison.

**Table S5.** Number of unique publications (accounting for multiple author positions on papers; data used for. Figure 1**) for indieSAGE/Vaccine Plus advocates and four control groups with n=16 authors.**

|  | Total | Research | Opinion | Review/Methods | Analysis |
| --- | --- | --- | --- | --- | --- |
| **2016-2019 (Pre-pandemic)** |  |  |  |  |  |
| IndieSAGE/Vaccines Plus | 87 | 2 | 74 | 0 | 11 |
| SAGE | 4 | 1 | 2 | 0 | 1 |
| GBD UK | 0 | 0 | 0 | 0 | 0 |
| UK most highly cited | 30 | 18 | 7 | 3 | 2 |
| UK most COVID-19 papers | 9 | 3 | 5 | 1 | 0 |
| **2020-2023 (Pandemic)** |  |  |  |  |  |
| IndieSAGE/Vaccines Plus | 356 | 17 | 310 | 7 | 22 |
| SAGE | 17 | 10 | 4 | 0 | 3 |
| GBD UK | 4 | 2 | 2 | 0 | 0 |
| UK most highly-cited | 23 | 6 | 14 | 2 | 1 |
| UK most COVID-19 papers | 27 | 15 | 8 | 3 | 1 |
| **2024 (Post-pandemic)** |  |  |  |  |  |
| IndieSAGE/Vaccines Plus | 16 | 0 | 16 | 0 | 0 |
| SAGE | 0 | 0 | 0 | 0 | 0 |
| GBD UK | 0 | 0 | 0 | 0 | 0 |
| UK most highly-cited | 4 | 0 | 3 | 0 | 1 |
| UK most COVID-19 papers | 1 | 1 | 0 | 0 | 0 |

**Table S6.**
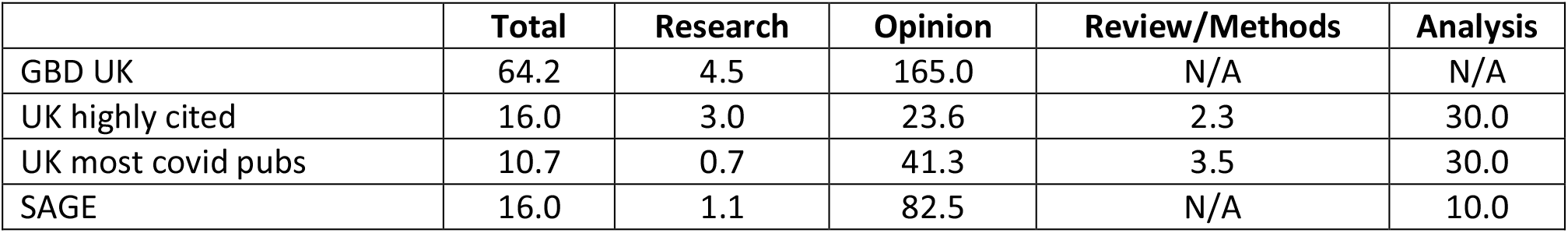
Ratios of author incidence, indieSAGE vs. other groups, including the BMJ-IndieSAGE freelance journalist (all papers in BMJ 2020-23)

|  | Total | Research | Opinion | Review/Methods | Analysis |
| --- | --- | --- | --- | --- | --- |
| GBD UK | 64.2 | 4.5 | 165.0 | N/A | N/A |
| UK highly cited | 16.0 | 3.0 | 23.6 | 2.3 | 30.0 |
| UK most covid pubs | 10.7 | 0.7 | 41.3 | 3.5 | 30.0 |
| SAGE | 16.0 | 1.1 | 82.5 | N/A | 10.0 |

**Table S7.** Ratios of author incidence, indieSAGE vs. other groups, excluding the BMJ-IndieSAGE freelance journalist (all papers in BMJ 2020-23)

|  | Total | Research | Opinion | Review/Methods | Analysis |
| --- | --- | --- | --- | --- | --- |
| GBD UK | 34.2 | 4.5 | 75.0 | N/A | N/A |
| UK highly cited | 8.5 | 3.0 | 10.7 | 2.3 | 30.0 |
| UK most covid pubs | 5.7 | 0.7 | 18.8 | 3.5 | 30.0 |
| SAGE | 8.5 | 1.1 | 37.5 | N/A | 10.0 |

**Table S8.**
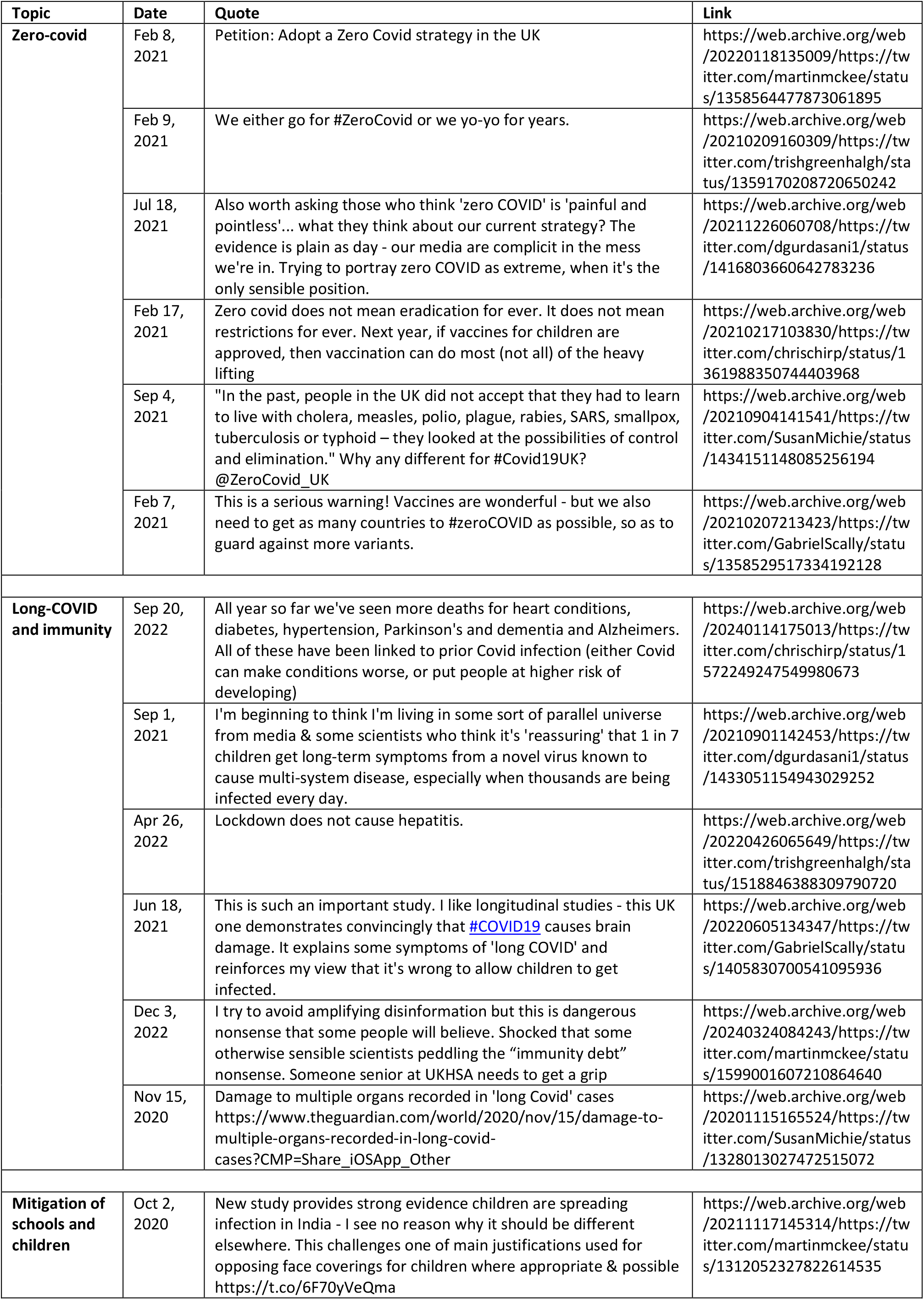
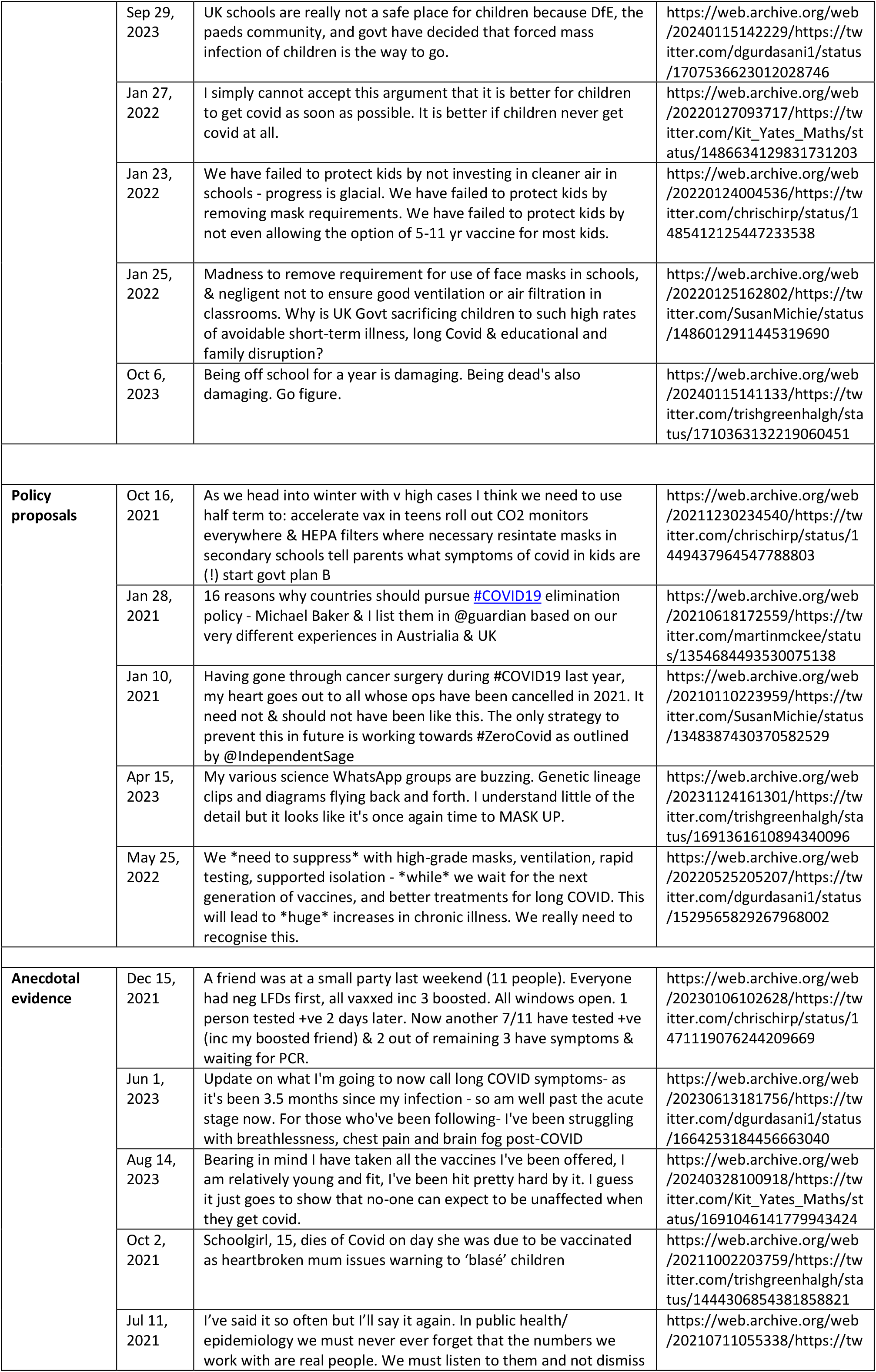
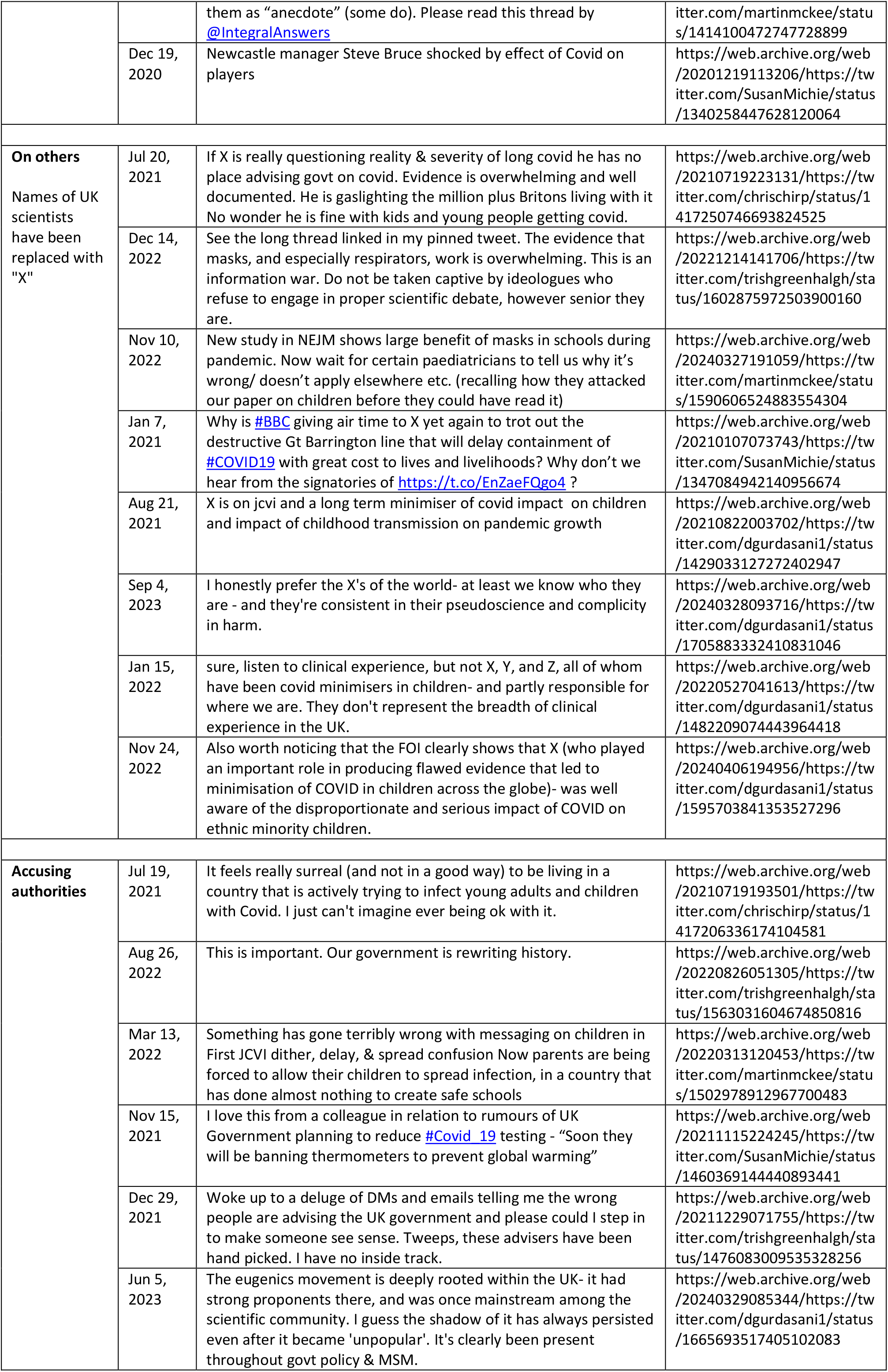
Selected views of indie-SAGE members from Twitter/X, 2020-2023.

